# A cross-sectional study of immune seroconversion to SARS-CoV-2 in front-line maternity health professionals

**DOI:** 10.1101/2020.06.24.20139352

**Authors:** Sohail Bampoe, Dominique Nuala Lucas, Georgina Neall, Penny Sceales, Reena Aggarwal, Kim Caulfield, Dimitrios Siassakos, Peter Mark Odor

## Abstract

COVID-19, the respiratory disease caused by the SARS-CoV-2, is thought to cause a milder illness in pregnancy with a greater proportion of asymptomatic carriers. This has important implications for the risk of patient-to-staff, staff-to-staff and staff-to patient transmission among health professionals in maternity. The aim of this study was to investigate the prevalence of previously undiagnosed SARS-CoV-2 infection in health professionals from two tertiary-level maternity units in London, UK and to determine associations between HCW characteristics, reported symptoms and serological evidence of prior SARS-CoV-2 infection.

200 anaesthetists, midwives and obstetricians with no previously confirmed diagnosis of COVID-19 were tested for immune seroconversion using laboratory IgG assays. Comprehensive symptom and medical histories were also collected. 5/40 (12.5%; 95% CI: 4.2-26.8) anaesthetists, 7/52 (13.5%; 95% CI: 5.6-25.8%) obstetricians and 17/108 (15.7%; 95% CI: 9.5-24.0%) midwives were seropositive, with an overall total of 29/200 (14.5%; 95% CI: 9.9-20.1%) of maternity healthcare workers testing positive for IgG antibodies against SARS-CoV-2. Of those who had seroconverted, 10/29 (35.5%) were completely asymptomatic. Fever or cough were only present in 6/29 (20.7%) and (10/29 (34.5%) respectively. Anosmia was the most common symptom occurring in 15/29 (51.7%) seropositive participants and was the only symptom that was predictive of positive seroconversion (OR 18; 95% CI 6 – 55). 58.6% of those who were seropositive had not self-isolated at any point and continued to provide patient care in the hospital setting.

This study was the largest study of baseline immune seroconversion in maternity healthcare workers conducted to date and reveals that 1 in 6 were seropositive, of whom 1 in 3 were asymptomatic. This has significant implications for the risk of occupational transmission of SARS-CoV-2 for both staff and patients in maternity and regular testing of staff, including asymptomatic staff should be considered to reduce transmission risk.

## Introduction

Serological testing to assess the extent of seroconversion in a population plays a vital role in containing an infectious outbreak. It can provide information on the prevalence of the infection in a cohort of people and also provide information about how a disease spreads. In the United Kingdom, current data from the Office of National Statistics (ONS) suggests that in May 2020, 6.78% of the population in England had detectable antibodies to SARS-CoV-2, the virus that causes COVID-19 [1]. A similar study of community seroconversion in Los Angeles, USA, reported a comparable seroconversion proportion of 4% [2]. A particular area of concern are people who seroconvert without having experienced clinical features of infection. A study of 215 pregnant women presenting for delivery in New York, USA, reported that 13.7% tested positive for active SARS-CoV-2 infection, 87.9% of whom were asymptomatic [3]. Although this study did not test for seroconversion, these data might suggest that the overall infection rate in the pregnant population may be much higher than the general population due to the presence of a higher proportion of asymptomatic carriers. Obstetric care has continued throughout the COVID-19 pandemic. Given that third-trimester pregnancy care involves multiple ongoing encounters with healthcare professionals and a potentially high proportion of reported asymptomatic carriers, obstetric healthcare professionals may be at an elevated risk of patient-to-staff, staff-to-staff, and also staff-to-patient transmission. Several studies have reported an elevated risk of transmission to, and seroconversion, in front-line non-obstetric healthcare workers (HCWs) in several settings, including in Wuhan, China, and in London, UK [4,5]. Only one single-centre study reports seroconversion rates in an obstetric unit, however this was following a COVID-19 outbreak amongst unit staff in that particular unit in Ohio, USA [6]. To date, no studies have reported a baseline seroconversion rate of obstetric health care workers providing front-line, patient-facing care during the current SARS-CoV-2 pandemic.

The primary aim of this study was to investigate the prevalence of previously undiagnosed SARS-CoV-2 infection in two tertiary level maternity units in London, UK. The secondary aim was to determine associations between HCW characteristics, reported symptoms and serological evidence of prior SARS-CoV-2 infection.

## Methods

Following approval from the NHS Health Research Authority (ref: 20/WA/0138), we recruited maternity healthcare workers from two, large central London teaching hospitals between 11^th^ May 2020 and 5^th^ June 2020. Included were midwives working on labour ward or post-natal care, anaesthetists or obstetricians, age ≥ 18 years. Participants were excluded if they had a history of SARS-Cov-2 infection diagnosed by PCR from nasopharyngeal samples, or if they were currently symptomatic for SARS-CoV-2 infection. Staff working in non-patient-facing roles were excluded. Members of the research team approached potential participants and all of those enrolled into the study provided written consent. Participation was voluntary.

Staff recruited into the study completed a questionnaire that captured demographic data, a self-reported medical history and a symptom history since January 2020. Upon completion of the questionnaire, the research team collected 10mls of blood in serum separating tubes (SST) from each participant. Blood samples were centrifuged for 10 minutes at 3100 rpm and separated serum was frozen at −80 °C ahead of transport to the laboratory.

Serological testing was performed using Abbott Laboratories’ (USA) two-step, chemiluminescent micro-particle immunoassay (CMIA) technology, approved by the United States Food and Drug Administration (FDA) and Public Health England (PHE). This assay detects the presence of IgG antibodies formed against the nucleocapsid protein antigen of SARS-CoV-2 in samples of patient serum [7]. Serum samples are combined with antigen-coated paramagnetic micro-particles and assay diluent before incubation. After a wash cycle, anti-human IgG acridinium-labelled conjugate is added and a further wash cycle is performed before the addition of trigger solutions. As a result of the addition of the trigger solutions, a chemiluminescent reaction occurs that is measured as a relative light unit (RLU). The RLU is measured by Abbott Laboratories’ proprietary system known as the ARCHITECT isystem. The RLU detected by the ARCHITECT system optics is directly related to the amount of IgG antibodies present[7]. The system also measures the RLU from a control sample. If dividing the sample RLU by the control RLU produces an index of <1.4, the sample is considered to be negative for SARS-CoV-2 IgG antibodies. An index >1.4 is considered to be a positive result [7]. This test has 100% sensitivity and 99.6% specificity as reported by the manufacturer and the US FDA [8].

All data collected through the questionnaire, in addition to results of serology testing were entered into a RedCap™ relational database system.

The required sample size was estimated by assuming a prevalence of seroconversion of 15%. Enrolling a sample size of 196 participants would allow precision of +/- 5% with confidence intervals of 95%. If the actual prevalence was lower than estimated, then the precision of our data sample would improve.

Statistical analysis was performed using SPSS (IBM, USA). Summary statistics were used to describe cohort demographics and symptom frequency. Frequency and percentage were presented for each categorical variable. For skewed continuous variables, the median and interquartile range (IQR) were calculated along with the range. The proportion of seroconversion were calculated with binomial 95% confidence intervals (CI).

Categorical variables were analysed using chi-squared or Fisher’s exact test. Binary logistic regression analysis was performed to determine factors associated with seroconversion, including age, gender, BMI, occupational role, ethnicity and the presence of recognised COVID-19 symptoms. The model’s calibration, or its fit to the data, was subsequently assessed with the Hosmer–Lemeshow test; this was determined by the degree of agreement between the risk score probabilities that had been predicted by the model, and the probabilities that were actually observed. Interaction effects were introduced to optimise model fit.

## Results

A total of 200 obstetric healthcare professionals were tested over a three-week period between May and June 2020. This included 40 (20.0%) anaesthetic physicians practising in maternity, 108 (54.0%) midwives, and 52 (26.0%) obstetric physicians. Baseline demographics are reported in Table 1.

**Table 1.** Participant demographic, occupational and comorbid characteristics. Values are n and range [IQR], standard deviation and proportion (%), as appropriate.

|  | Participants (n = 200) |
| --- | --- |
| Age (y) | 37 (30 – 37 [21 – 66]) |
| Weight (kg) | 69.4 (13.4) |
| BMI (kg m<sup>-2</sup>) | 25.1 (5.0) |
| Female | 167 (83.5) |
| Occupation |  |
| Anaesthetist | 40 (20.0%) |
| Midwife | 108 (54.0%) |
| Obstetrician | 52 (26.0%) |
| Ethnicity |  |
| Caucasian | 139 (69.5%) |
| BAME | 61 (30.5%) |
| Asian or Asian British | 25 (12.5%) |
| Black or Black British | 25 (12.5%) |
| Mixed | 9 (4.5%) |
| Other | 2 (1.0%) |
| Smoker |  |
| Never smoked | 141 (70.5%) |
| Former smoker | 36 (18.0%) |
| Smoker | 13 (6.5%) |
| Unknown | 10 (5.0%) |
| Comorbidities |  |
| Immunosuppressed or receiving steroids | 0 |
| Cardiac disease | 0 |
| Hypertension | 10 (5.0%) |
| COPD | 0 |
| Diabetes | 1 (0.5%) |
| Diabetes with end organ complications | 0 |
| Asthma | 21 (10.5%) |
| Chronic kidney disease | 0 |
| Moderate/severe hepatic disease | 1 (0.5%) |

IgG antibodies to SARS-CoV-2 were detected in 29/200 (14.5%; 95% CI: 9.9-20.1%) of obstetric HCW. Seroconversion amongst individual staff groups was: midwifery 17/108 (15.7%; 95% CI: 9.5-24.0%); anaesthesia 5/40 (12.5%; 95% CI: 4.2-26.8); obstetrics 7/52 (13.5%; 95% CI: 5.6-25.8%). Participants were predominantly female (83.5%). Seroconversion proportions were similar at both hospitals.

Of the seroconverted staff members 10/29 (34.5%) were completely asymptomatic. In those with a history of symptoms and who seroconverted, the most common symptoms were anosmia 15/29 (51.7%) and headache. 10/29 of the seropositive participants (34.5%) reported a history of dry or productive cough and 6/29 (20.7%) reported a history of fever.

Twelve out of 29 (41.4%) seropositive participants had self-isolated at least once during the pandemic and 17/29 (58.6%) continued to work throughout either because they were asymptomatic or their symptoms did not qualify them at the time for self-isolation (10 and 7 members of staff respectively). Logistic regression found that the presence of anosmia was the only significant predictor of antibody seroconversion (OR 18; 95% CI 6 – 55). No other demographic or symptom variables were predictive of immune seroconversion including age, staff group, BMI, ethnicity, fever or cough (Table 3).

**Table 2.** Reported symptoms and SARS-CoV-2 detection by serology.

| Symptoms | Seroconverted<br>(n = 29) | No antibodies<br>detected<br>(n =171) | p |
| --- | --- | --- | --- |
| Asymptomatic | 10 (34.5%) | 59 (34.5%) | 0.99 |
| Dry cough or fever or anosmia | 16 (55.2%) | 53 (31.0%) | 0.11 |
| Dry cough | 10 (34.5%) | 41 (24.0%) | 0.23 |
| Fever | 6 (20.7%) | 15 (8.8%) | 0.05 |
| Anosmia | 15 (51.7%) | 9 (5.3%) | <0.01 |
| Headache | 14 (48.3%) | 61 (35.7%) | 0.19 |
| Myalgia | 12 (41.4%) | 42 (24.6%) | 0.59 |
| Nasal congestion | 6 (20.7%) | 25 (14.6%) | 0.40 |
| Sore throat | 6 (20.7%) | 52 (30.4%) | 0.28 |
| Shortness of breath | 5 (17.2%) | 24 (14.0%) | 0.65 |
| Diarrhoea | 4 (13.8%) | 24 (14.0%) | 0.97 |
| Nausea or vomiting | 3 (10.3%) | 14 (8.2%) | 0.70 |
| Photophobia | 2 (6.9%) | 0 (0%) | 0.02 |
| Productive cough | 1 (3.4%) | 9 (5.3%) | 0.68 |
| Abdominal pain | 1 (3.4%) | 12 (7.0%) | 0.47 |

**Table 3.** Factors associated with detection of antibodies to SARS-CoV-2 in maternity healthcare workers. Values are OR (95% CI).

| Characteristic | OR (95% CI) | p |
| --- | --- | --- |
| Age |  |  |
| 18-30 | 1.0 (ref) |  |
| 31-50 | 1.1 (0.3-4.0) | 0.92 |
| 51-70 | 1.1 (0.2-5.4) | 0.87 |
| Obesity | 0.6 (0.1-2.6) | 0.49 |
| Gender |  |  |
| Female | 1.0 (ref) |  |
| Male | 1.1 (0.2-5.5) | 0.88 |
| Ethnicity |  |  |
| Caucasian | 1.0 (ref) |  |
| BAME | 1.6 (0.6-4.5) | 0.36 |
| Fever | 1.5 (0.3 – 7.7) | 0.61 |
| Anosmia | 18.2 (6.0-55.2) | <0.01 |
| Headache | 1.1 (0.4-3.4) | 0.86 |
| Self-isolated | 0.4 (0.1-1.9) | 0.28 |
| Dry cough | 1.0 (0.3 – 3.4) | 0.86 |

## Discussion

This is the largest study to report baseline immune seroconversion to SARS-CoV-2 in obstetric healthcare workers to date. We found that one out of every six front-line, obstetric healthcare workers had been infected with SARS-CoV-2 as revealed by the presence of IgG antibodies. Seroconversion was more than twice as prevalent in UK obstetric HCWs as in the general population in the UK and three times as prevalent as in the general population in the USA [1,2]. However, the prevalence of seroconversion of HCWs in these London hospitals was similar to estimates of seroconversion in the general population in Greater London at 14.5%. based on sampling from blood donors [9].

Although these results would suggest that obstetric HCWs are at a similar risk of exposure to COVID-19 as the general population, the prevalence of seroconversion among this staff group appears to be lower than reported in other frontline HCW groups. Houlihan et al. measured seroconversion in 200 HCW in London working in the emergency department, acute medical unit, COVID-19 cohort wards, intensive care unit and haematology wards. They used serial serology tests over three months and reported a collective seroconversion proportion of 45.3% [5]. One possible explanation for the difference in seroconversion rates between obstetric HCWs and other front-line HCWs could be due to the transmission characteristics of SARS-CoV-2. The transmission characteristics of SARS-CoV-2 appear to be similar to those of influenza in that viral shedding appears to occur in those who are asymptomatic and also in the pre-symptomatic phase of infection [10]. In patients infected with influenza, quantitative viral load in asymptomatic individuals is less in upper respiratory tract secretions and the duration of viral shedding is also shorter than in symptomatic individuals [11]. Therefore, if the above assumptions are correct, then it may be reasonable to extrapolate that asymptomatic individuals infected with SARS-CoV-2 pose a lower transmission risk to others. Hence the reported greater proportion of asymptomatic infection in pregnancy and generally milder symptom profile in pregnant patients may explain why obstetric HCWs appear to have a lower risk of occupational viral transmission, as SARS-CoV-2 pregnant patients may be less infective than respiratory or acute emergency patients [3,12].

The risk posed by infectious staff members to colleagues, pregnant women, and their offspring is unknown. At the beginning of the current SARS-CoV 2 pandemic, the UK government advised all those with a persistent cough or fever >37.8°C to self-isolate. Our data revealed that only 41.4% of HCW who were seropositive met those criteria and self-isolated at any point. This means that 58.6% continued to work and commute despite active SARS-CoV-2 infection. Indeed, our data showed that in this population, neither cough nor fever predicted seropositivity and the only symptom predictive of immune seroconversion was anosmia. The UK government have since added anosmia as a symptom that mandates self-isolation. Our study strongly supports this updated advice, albeit it would still fail to isolate about 6 out of 10 infected staff members. Of importance, it was not until 15^th^ June 2020 that all staff members in patient-facing areas were advised by Public Health England to wear surgical masks to reduce the risk of infection to others [13].

In maternity settings, SARS-CoV-2 poses risks to staff and women alike, but also potentially to their unborn or born babies. The United Kingdom Obstetric Surveillance System (UKOSS) collected data from 427 pregnant women admitted with confirmed SARS-CoV-2 infection and reported an estimated incidence of maternal hospitalisation with COVID-19 of 4.9 per 1000 pregnancies [12]. The majority of patients were in the second and third trimesters of pregnancy and suffered mild disease. In this study, of 266 completed maternities there were three stillbirths, with two potentially related to COVID-19. With current UK stillbirth rates about 4 in 1000, only one stillbirth would have been expected in the UKOSS cohort [13, 14]. Studies of placentas from COVID-19 infected women have highlighted the potential of the wider adverse impact of COVID-19 infection on pregnancy [15]. Moreover, even if the incidence of serious neonatal COVID-19 infection is low, possible vertical or perinatal transmission has been reported and the issue of separation of babies from infected mothers has been debated [16,17]. Transmission to the newborn by infectious asymptomatic members of staff present at birth, or to their mothers by asymptomatic staff members has not been investigated to date. Knowledge of the incidence of symptomatic and asymptomatic seroconversion of maternity staff and how it compares to the general population, to staff in other healthcare settings, and to maternity patients, is useful for providing context to such investigations, particularly in case of future additional waves of infection.

Until we have robust evidence as to the risk posed by asymptomatic infected individuals to others, and as to the risk of COVID-19 to babies, particularly in utero, our study suggests that extreme caution is advisable in maternity settings, particularly the consistent use of effective PPE. We also recommend that all obstetric healthcare institutions should consider regular serology testing for staff, as well as the immediate isolation of any staff who develop anosmia, even in the absence of cough or fever. Regular testing and consistent use of PPE are likely to be the cornerstones of pandemic control.

## Strengths and limitations

Our study used antibody seroconversion as a marker of previous COVID-19 infection. Based upon measurements in other similar viruses (REF), the duration of antibody titres sufficient to be detected is likely to be for at least six months. However, the exact duration of SARS-CoV-2 IgG antibody persistence remains unknown and recent studies have shown that mild cases might end up seronegative and only exhibit mucosal IgA [18]. As such it possible that some individuals who were infected earlier in the year, as well as some mild cases, did not exhibit sufficient antibody response at the date of testing. Our testing methodology may therefore underestimate the true seroconversion prevalence.

## Data Availability

Data available on request

## Author Contributions

Study conception: SB, PO, NL, DS

Data collection: SB, PO, GN, PS, RA, KC Manuscript preparation: SB, PO

Manuscript revision: SB, NL, DS, PO, GN, PS, RA, KC

